# Stability of influenza viruses in the milk of cows and sheep

**DOI:** 10.1101/2025.05.28.25328508

**Authors:** Jenna Schafers, Caroline J. Warren, Jiayun Yang, Junsen Zhang, Sarah J. Cole, Jayne Cooper, Karolina Drewek, Natalie McGinn, Mehnaz Qureshi, Scott M. Reid, Nunticha Pankaew, Wenfang Spring Tan, Sarah K. Walsh, Ashley C. Banyard, Ian Brown, Paul Digard, Munir Iqbal, Joe James, Thomas P. Peacock, Edward Hutchinson

## Abstract

In late 2023, H5N1 high pathogenicity avian influenza (HPAIV) started circulating in dairy cattle in the USA. High viral titres were detected in milk from infected cows, raising concerns about onwards human infections. Although pasteurisation was shown to effectively inactivate influenza viruses in milk, unpasteurised milk still poses a risk of infection, both from occupational exposure in dairies and from the consumption of raw milk. We therefore assessed how long influenza viruses could remain infectious for in milk without heat inactivation. We examined the stability of a panel of influenza viruses in milk, including a contemporary H5N1 HPAIV and a variety of other influenza A and D viruses. We incubated viruses in cows’ milk under laboratory conditions: at room temperature to simulate exposure in dairies and at 4°C to simulate exposure to refrigerated raw milk. Following an isolated report of H5N1 viral RNA being detected in milk from a sheep in the UK, we also carried out similar experiments with a laboratory strain of IAV in sheep’s milk. Although the survival of influenza viruses in milk was variable, we consistently found that under laboratory conditions substantial viral infectivity remained over periods when people might reasonably be exposed to infected milk – for over a day at room temperature and for more than 7 days when refrigerated. Our results highlight the zoonotic risk of H5N1 HPAIV in raw milk from infected animals and reinforce the importance of taking measures to mitigate this risk.

## Introduction

In March 2024, following reports of significantly reduced milk yield, H5N1 high pathogenicity avian influenza virus (HPAIV) was detected circulating among dairy cattle in the United States of America (USA). The virus soon became widely distributed in dairy cattle across the country, and subsequent phylogenetic analysis suggested that the first introduction into cattle had occurred in late 2023 [1]. The outbreak was unexpected, as although cattle were known to be susceptible to experimental infection with influenza A viruses (IAVs), the genus to which HPAIVs belong, they were not considered to be a species that was generally affected by these viruses[2]. Also unexpected was the mode of transmission – while influenza viruses in mammals are primarily thought of as respiratory infections, in cattle the virus was shed at extremely high levels in milk [3-6]. At the time of writing, H5N1 HPAIV has been detected in over a thousand herds of cattle across the contiguous USA, as well as in dozens of infected dairy workers and in several cryptic infections of people reporting no direct contact with infected animals[6]. The genetic material of H5N1 HPAIV has been detected in consumer dairy products in across the USA, with approximately 20% of retail milk samples testing positive for viral RNA in some affected areas [7]. Although studies have confirmed that pasteurisation effectively inactivates H5N1 HPAIV in milk [8-17], unpasteurised (‘raw’) milk may pose an infection risk, through both occupational exposure and consumption.

Occupational exposure to H5N1 contaminated milk is a risk for dairy workers in affected areas, and one study has reported H5 seroprevalence of 7% in dairy workers in such settings [18]. Multiple human infections have now been associated with dairy work in the USA[6], such as a dairy worker who developed unilateral conjunctivitis following a splash of milk into the eye, acquired during milking without the use of eye or face protection, and was subsequently diagnosed as being infected with H5N1 HPAIV from cattle [19].

The risks posed by consuming raw milk, or milk products, that are contaminated with H5N1 are harder to assess. The virus can remain infectious in milk and in some milk products: adding influenza viruses to milk only marginally reduces their infectivity in *in vitro* assays [8], and the virus appears to retain its infectivity for over 60 days in cheese made from unpasteurised cow’s milk [20]. Consumption of H5N1 contaminated milk has been demonstrated to be a direct infection mechanism in animals. Over 50% of cats on a Texas dairy farm fell ill after consuming raw colostrum and milk from cows infected with H5N1 [3] and, in experimental studies, H5N1-contaminated milk was infectious through direct oral inoculation of mice and was a plausible route of transmission between lactating mice or ferrets and their pups [16, 21, 22]. However, it should be noted that the specific details of how different animals eat, and of how experimental infections are undertaken, mean that none of these studies are an ideal proxy for human consumption of milk or dairy products. At the current time the infectious dose of H5N1 through the consumption of milk by any animal, including humans, remains unknown. What we do know is that milk from H5N1-infected cattle can contain virus at extremely high infectious titres [3, 23], and repeated observations in the USA of human infections by bovine H5N1 for people without a clear history of exposure to infected animals suggests that consumer exposure to H5N1 in raw milk may pose a meaningful risk of infection.

Influenza virus particles lose their infectivity over time and, given that unpasteurised (‘raw’) milk could contain H5N1 influenza virus, it was important to determine how long contaminated milk could remain infectious. The likely conditions of influenza virus contaminated milk differ in the two scenarios outlined above. In the dairy industry, H5N1 HPAIV might be found in milk present on surfaces across dairies, including milking and milk transport equipment (upstream of pasteurisation, if this is carried out). This could pose a transmission risk to dairy farm workers by direct contact or through aerosol exposure during milking, transport or cleaning [24]. In these settings, spilt milk is likely to remain at ambient temperatures until it is removed by cleaning. During this time, it may dry out and be exposed to a wide variety of environmental contaminants and to ultraviolet light. In contrast, consumers of raw milk are more likely to be exposed to milk that has been refrigerated, is largely free of environmental contaminants and which is potentially stored away from ultraviolet light sources, both at the point of purchase and in domestic refrigerators.

In this paper, we assessed the risk that influenza viruses could remain stable in unpasteurised milk. For a panel of influenza viruses, including H5N1 HPAIV, we assessed how rapidly infectivity was lost in cow’s milk at both room temperature (simulating dairy farm environments) and 4°C (simulating customer refrigeration). In response to a recent isolated case of a lactating sheep in the UK shedding H5N1 viral RNA in its milk [25], we also assessed the stability of an influenza virus in sheep’s milk. Our findings show that, in the absence of other inactivating factors such as ultraviolet light, detergents or desiccation, H5N1 and other influenza A viruses can remain infectious in milk for over a day at room temperature and more than 7 days at 4°C – essentially, for the longest period over which milk might plausibly be left in each setting before being disposed of. Our results suggest that unpasteurised milk, whether present on surfaces in dairies or sold for human or animal consumption, could serve as a source of H5N1 HPAIV exposure in areas where this virus is being shed by infected cows or other dairy animals.

## Results

To assess the survival of H5N1 HPAIV in milk, we initially tested the wild-type strain A/chicken/Scotland/054477/2021 (AIV09/AB genotype) under SAPO containment level 4 conditions. The virus was mixed at a ratio of 1:10 (v/v) with unpasteurised (‘raw’) milk. It was incubated in closed tubes, to prevent evaporation, which were stored within sealed polystyrene boxes, to stabilise the temperature and to prevent UV light inactivation. The mixture was held at either room temperature (∼20°C) or chilled (∼4°C) for up to 12 days. In parallel, virus was mixed 1:10 with phosphate-buffered saline (PBS) and stored under identical conditions. Viral infectivity was assessed using TCID_50_ assays. The H5N1 virus slowly lost infectivity over time in milk, at room temperature and when refrigerated. Under either condition, infectious titres remained detectable in raw milk for over 7 days (Figure 1).

**Figure 1.**
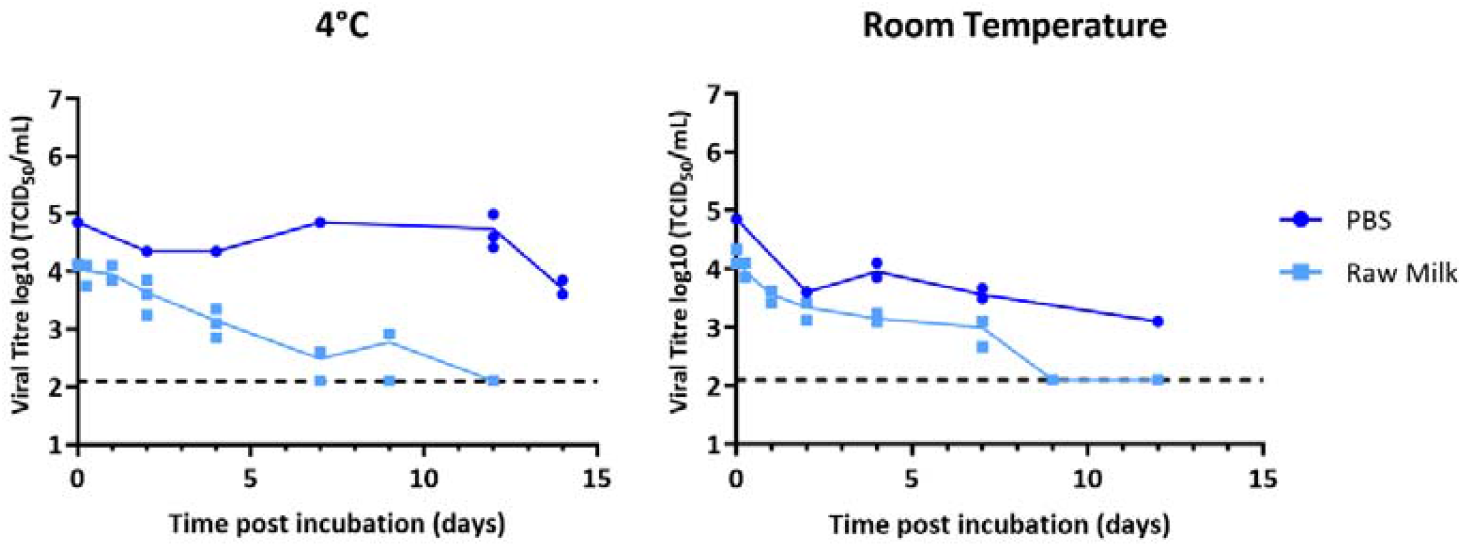
Stability of H5N1 HPAIV in milk. H5N1 HPAIV was mixed with raw cow’s milk or PBS and either incubated at room temperature (∼20°C) or chilled (4°C) for the indicated times. Infectivity was measured by TCID_50_. Data points show three independent repeats, with lines connecting the mean values. Limit of detection (LoD) = 126 TCID_50_/mL.

To assess whether this stability was a consistent property of other influenza viruses, we tested a panel of additional strains (Table 1). This included a reassortant influenza virus we had generated by reverse genetics which carried the internal genes of the laboratory strain A/Puerto Rico/8/1934 (PR8) and the external genes of a bovine H5N1 clade 2.3.3.4b (with the HA ‘de-engineered’ to replace the multi-basic cleavage site, which is associated with high pathogenicity, with a single-basic cleavage site). In addition, it included three variants of PR8 which were handled in different laboratories, two low pathogenicity avian influenza A viruses (LPAIVs) and an influenza D virus that naturally infects cattle. These samples were incubated in cow’s milk (pasteurised whole milk purchased in the UK) or buffered media (PBS or Dulbecco’s Modified Eagle Medium (DMEM)) under the same conditions described above, and their infectivity measured by plaque assay. We observed substantial variation in viral stability, particularly at room temperature, where infectivity in milk could be detected for more than 11 days in some cases but reached the limit of detection within 2 days in others. At 4°C, most viruses remained infectious for over 7 days, and in some cases for considerably longer (Figure 2).

**Table 1.** Influenza viruses used in the study.

| Short Name | Strain name and details |
| --- | --- |
| H5N1 | A/chicken/Scotland/054477/2021 (H5N1), a high pathogenicity avian influenza virus |
| PR8 (PR8-PI, PR8-RI, PR8-BF) | A/Puerto Rico/8/1934 (H1N1), a laboratory-adapted influenza A virus. PR8-RI refers to data collected at the Roslin Institute, and PR8-PI to data collected at the Pirbright Institute. |
|  | PR8-BF refers to 'BrightFlu,' a PR8 derivative encoding a fluorescent marker [26], for which data were collected at the MRC-University of Glasgow Centre for Virus Research |
| PR8-Cattle | A reassortant virus with the internal gene of A/Puerto Rico/8/1934 (H1N1), and the HA, NA gene of A/dairy cow/Texas/24-008749001-original/2024 (H5N1) |
| H5N2 | A/wild-duck/Italy/17VIR69261/2017 (H5N2), a low pathogenicity avian influenza virus |
| H5N3 | A/Duck/Singapore/97 (H5N3), a low pathogenicity avian influenza virus |
| IDV | D/bovine/France/5920/2014, a separate genus of influenza virus that that naturally infects cattle |

**Figure 2.**
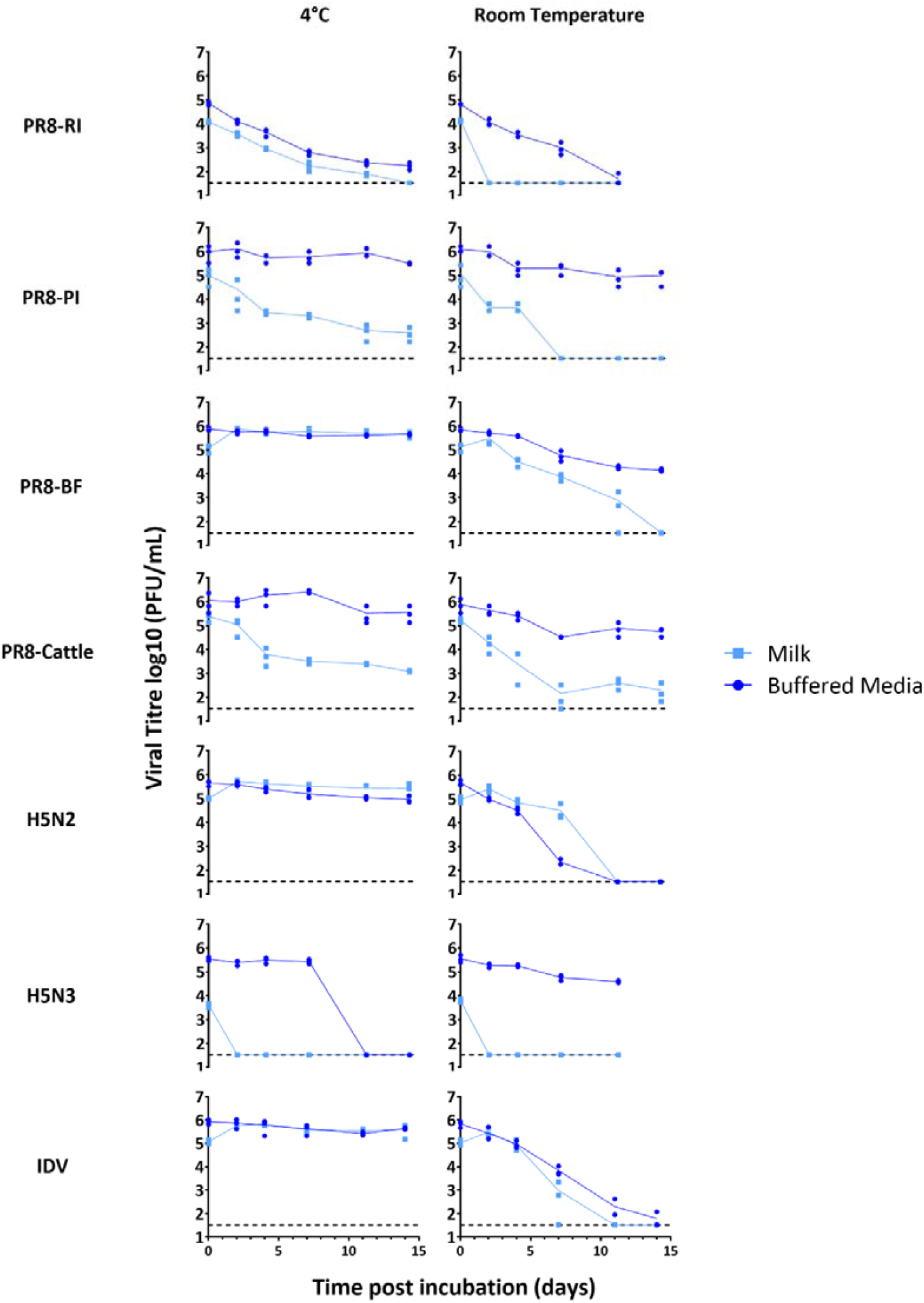
Stability of a panel of influenza viruses in milk. Viruses were mixed with pasteurised whole cow’s milk or buffered media and either incubated at room temperature (∼20°C) or chilled (4°C) for the indicated times. Infectivity was measured by plaque assay. Data points show three independent repeats (each point is the mean of duplicate technical replicates, except for IDV, which was measured once), with lines connecting the mean values. Limit of detection (LoD) = 33 PFU/mL.

We investigated whether the input viral titre had influenced the decay rate by testing a stock of PR8 that had been mixed with pasteurised milk to give final titres ranging from 10^4^ to 10^6^ PFU/mL. The infectivity decay rate was not obviously affected by the starting titre (Figure 3), although as expected, and consistently with the data in Figure 2, higher titre samples took longer to reach the limit of detection.

**Figure 3.**
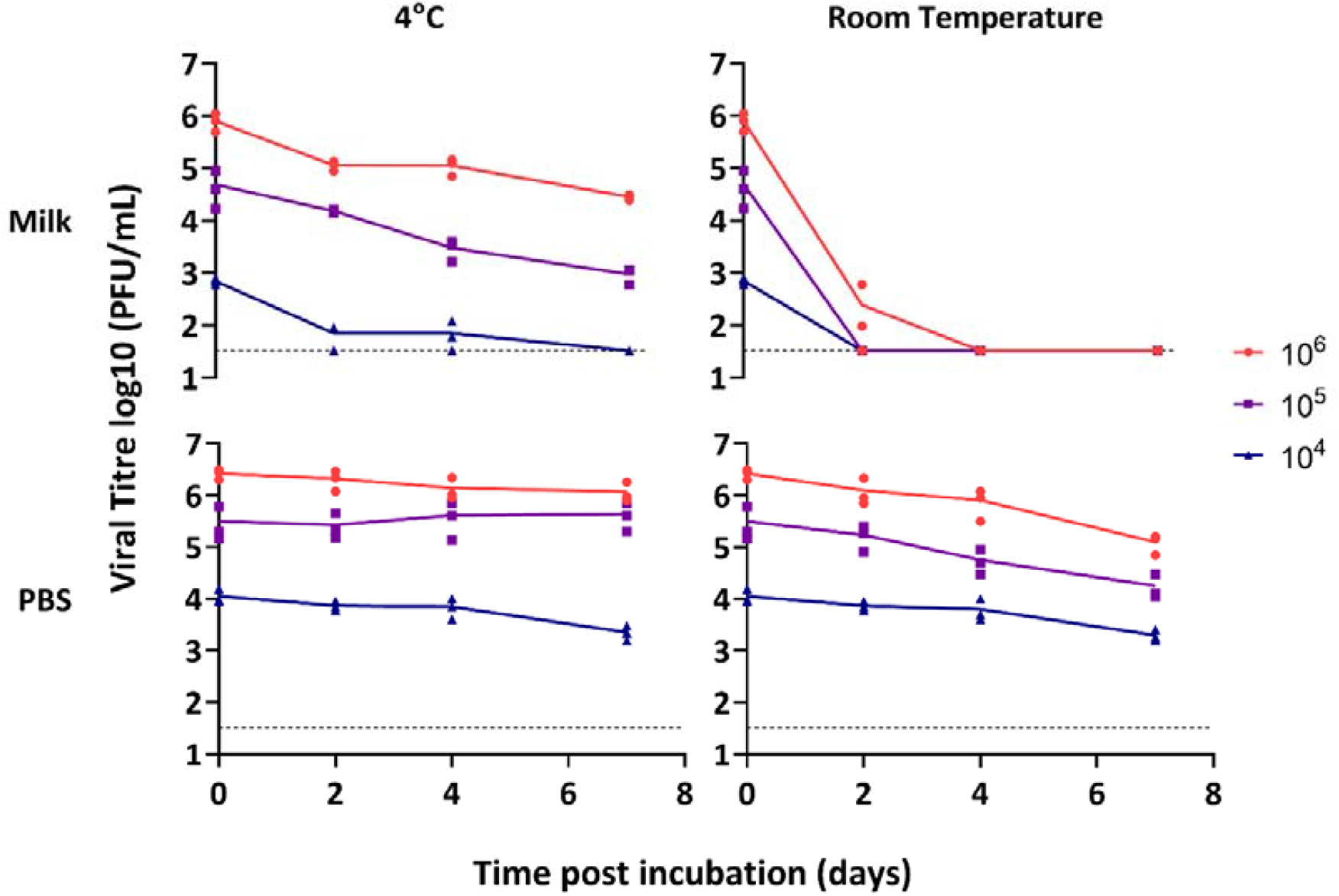
Stability of influenza virus in whole milk at different input viral titres. PR8-BF was diluted in DMEM, then mixed 1:10 with pasteurised whole cow’s milk to give the final titres shown and either incubated at room temperature (∼20°C) or chilled (4°C) for the indicated times. Infectivity was measured by plaque assay. Data points show three independent repeats, with lines connecting the mean values. Limit of detection (LoD) = 33 PFU/mL.

Finally, in response to an isolated case in the UK in which milk from a lactating sheep was found to be positive for H5N1 HPAIV viral RNA [25], we tested viral stability in sheep’s milk. In sheep’s milk, PR8-BF remained infectious for more than 7 days at 4°C and up to 4 days at room temperature (Figure 4), suggesting that its stability in sheep’s milk was broadly similar to that in cow’s milk.

**Figure 4.**
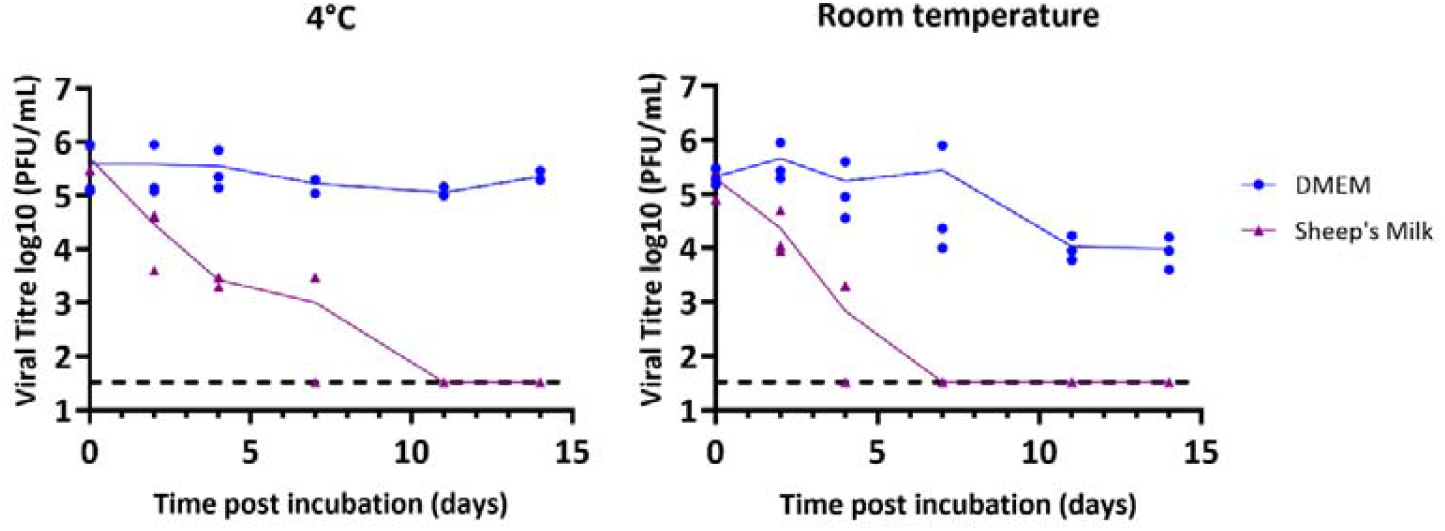
Stability of influenza virus in sheep’s milk. PR8-BF was mixed with pasteurised whole sheep’s milk, or with DMEM, and either incubated at room temperature (∼20°C) or chilled (4°C) for the indicated times. Infectivity was measured by plaque assay. Data points show three independent repeats, with lines connecting the mean values. Limit of detection (LoD) = 33 PFU/mL.

Taking our results together, the variation we observed between samples meant that we could not precisely determine a decay rate for influenza viruses in milk at either of the temperatures we tested. However, our main objective was to determine whether, in the absence of other inactivating factors such as desiccation, ultraviolet irradiation or environmental contaminants, influenza viruses could remain infectious in milk for long enough to pose a plausible infection risk to humans, and in this respect our data are very clear. Unless otherwise inactivated, influenza viruses can remain infectious in milk for over a day at room temperature (∼20°C) and for 7 days or more when refrigerated (4°C). These results, which were observed across a wide range of influenza strains, and tested in milk from diverse sources, cover the time periods over which humans are likely to be exposed to infected milk in either an occupational or a consumer setting.

## Discussion

In response to the detection of H5N1 HPAIV in milk from infected dairy cattle in the USA, we investigated the stability of influenza viruses in milk. We considered two scenarios – milk spilt in a dairy and left at ambient temperatures prior to cleaning, and raw milk refrigerated prior to and following sale to consumers. We tested the thermal stability of virus particles in milk under laboratory conditions, recognising that, particularly in the case of a dairy, a large number of additional environmental factors could inactivate viruses to a variable and unpredictable degree. These include (though are not limited to) ultraviolet irradiation, dehydration, changes in pH due to bacterial growth in the milk, adsorption to surfaces, and the introduction of substances including detergents and the residues of cleaning products. Our experimental design deliberately excluded these non-thermal factors. We designed our experiments by mixing viruses with milk in small, consistent, volumes, in sealed containers stored in the dark. Other than for the data shown in Figure 1, we attempted to reduce the effect of microbial contamination by using pre-pasteurised milk. Our experiments aimed to model the ‘worst case scenario’ for the persistence of viral infectivity in milk and should be seen as providing an upper-bound estimate for viral survival under real-world conditions.

Under our experimental conditions, H5N1 HPAIV was less stable in milk than in buffered media and had slightly greater stability in milk at 4°C than at room temperature. At both temperatures, however, viral infectivity remained above the limit of detection for over 7 days (Figure 1). To assess whether this rate of decay was consistent across different influenza virus strains, we tested additional influenza viruses under similar conditions. The results revealed substantial variation in viral stability across experiments (Figure 2), which had no obvious relation to the choice of virus strain, variation in the input titre of virus, or the matrix the inocula was added to for survival analysis (Figures 2 and 3).

Comparison with other studies of the stability of influenza viruses in milk [15, 27-30] further highlighted this variability (Table 2). In the current study, additional experiments using the PR8 and H5N3 viruses, in the same experimental setting but with different types of milk (one pre-pasteurised and one not), gave variable results, suggesting that the milk itself could introduce variation into the experiments (Figure S1). Milk is a non-standardised biological medium, whose composition is influenced by differences both within and between herds, as well as by environmental conditions once harvested [31]. Even after pasteurisation, milk remains non-sterile and undergoes pH changes over time due to bacterial fermentation and lactic acid production, which likely impact virus-milk interactions [31]. All of these factors limit our ability to predict a precise rate of decay of infectivity for influenza viruses in milk.

**Table 2.**
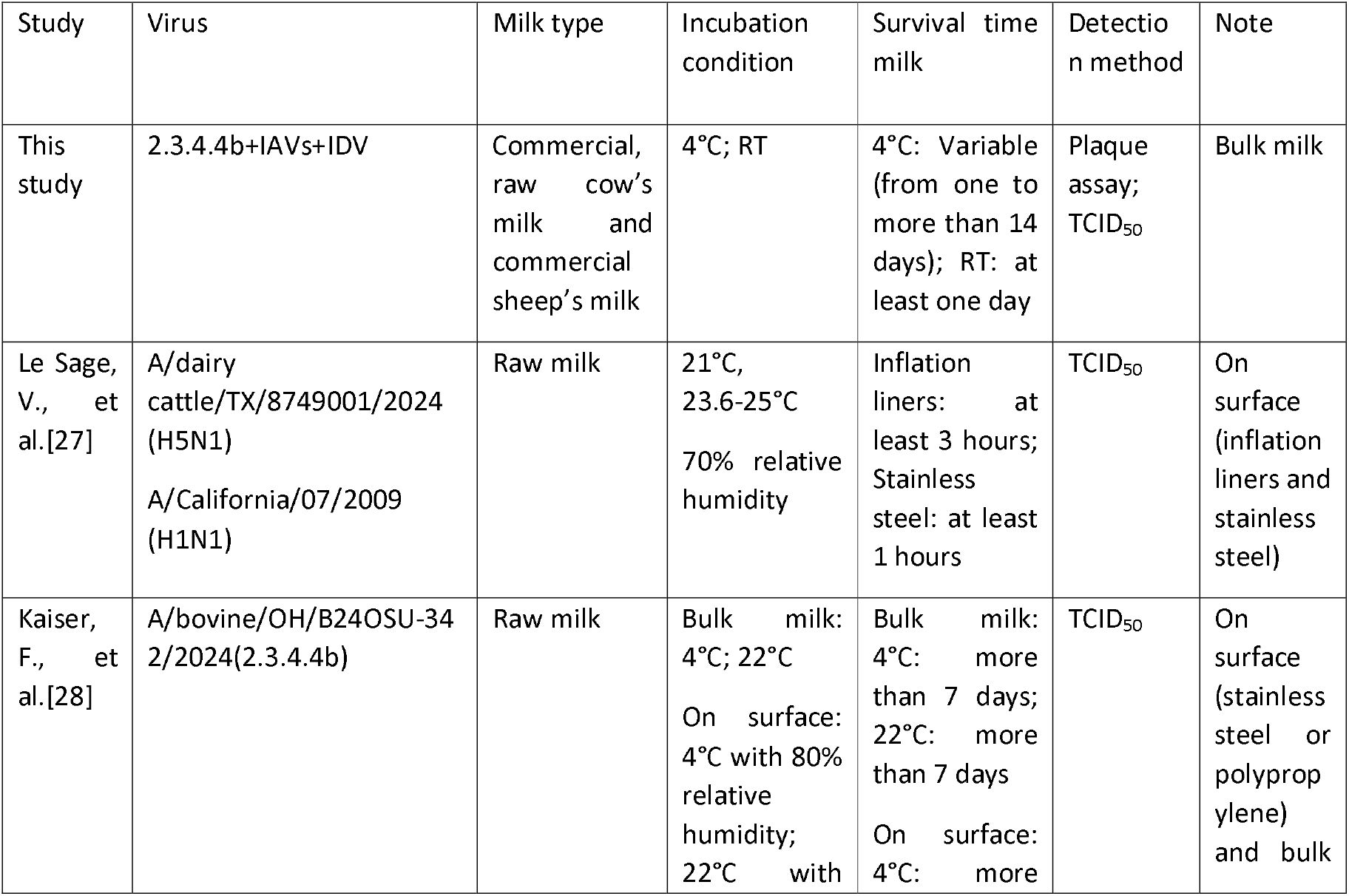
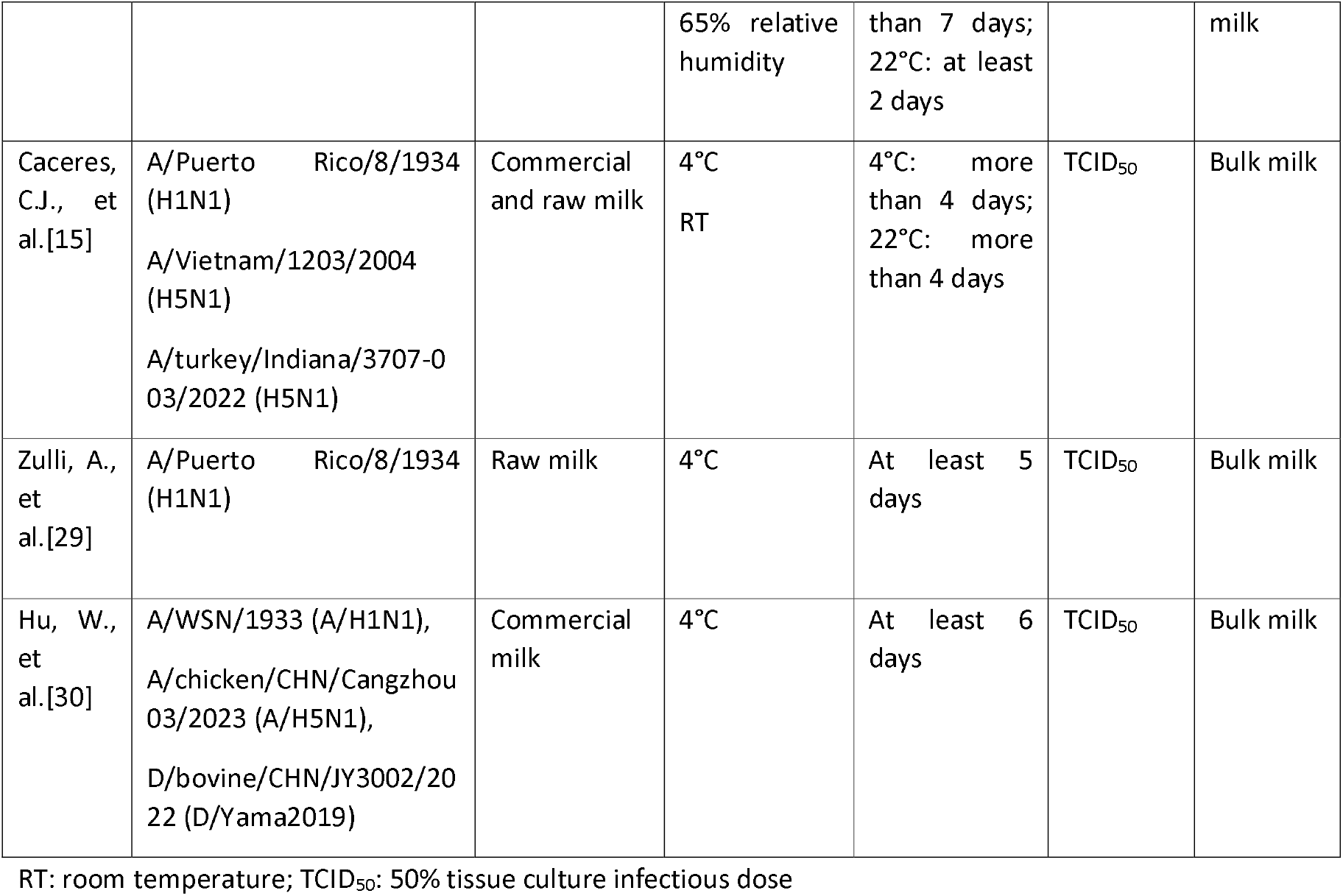
Comparison of studies of influenza virus stability in milk.

However, predicting a precise rate of decay is not ultimately necessary for a risk assessment of how long contaminated milk should be viewed as potentially retaining infectivity. Our findings consistently demonstrate that most influenza viruses can remain infectious in milk for longer than the milk itself is likely to remain in the environment – for over a day at room temperature, and for over a week at 4°C.

These findings underscore the potential infection risks associated with milk in areas where dairy animals are infected with H5N1 influenza viruses (at the time of writing, H5N1 is widespread in dairy cattle across the continental USA but has not yet been detected in cattle in other countries [32], and the detection of H5N1 in a sheep in the UK remains an isolated incident). Importantly, pasteurisation effectively inactivates influenza viruses [9, 13], however our data show that without pasteurisation influenza viruses can remain infectious for an extended period in milk. This creates potential occupational and consumer infection risks due to spilled milk in dairy environments, as well as the consumption, storage and transport of refrigerated, unpasteurised milk. In real-world scenarios environmental factors may reduce virus stability to an unpredictable extent. However, we also note that neither our study nor most of the other studies that have been performed clearly establish an upper limit for how long influenza viruses can remain infectious (Table 2). Indeed, recent work suggests that in unpasteurised cheese H5N1 influenza virus can remain measurably infectious for over 60 days [20]. In terms of spilled or stored milk, given the extremely high titres of virus that have been reported in milk from infected cattle [3, 23], even small volumes of milk in which the virus decays at the rates we observe could pose a significant infection risk for as long as they are likely to be encountered in the environment. Our data therefore suggest that the potential of infection from contaminated milk should be considered when planning control measures for H5N1 influenza in dairy settings, during milk processing and where raw milk is sold to consumers.

## Methods

### Cells and viruses

Experiments using A/Puerto Rico/8/1934 (H1N1; ‘PR8’) strain and its derivatives were conducted under biosafety level 2 (BSL-2) conditions. The PR8 derivative virus BrightFlu (PR8-BF), was generated by reverse genetics in 293 T cells as previously described [26, 33]. The PR8 derivative virus PR8-Cattle was a reassortant of PR8 with HA and NA genes from A/dairy cattle/Texas/24-008749-001-original/2024 (H5N1) (GISAID accession EPI_ISL_19014384). To permit handling of this virus under BSL-2 conditions, the polybasic cleavage site of the H5 HA segment was modified to a monobasic site. The HA and NA genes were then synthesised by GenScript and inserted into the pHW2000 vector. Virus rescue, using PR8 genes for the remaining 6 segments, was performed using the eight-plasmid bidirectional pHW2000 reverse genetics system in 293 T cells as previously described [34]. To produce working stocks, PR8 and PR8-BF were propagated on Madin-Darby Canine Kidney (MDCK) carcinoma cells, and PR8-Cattle was propagated in 9–10-day-old embryonated chicken eggs.

Additional viruses handled at BSL-2 were A/Duck/Singapore/97 (H5N3) (provided by Prof. Wendy Barclay, Imperial College London), A/wild duck/Italy/17VIR6926-1/2017 (H5N2) (from Dr. Isabella Monne, Istituto Zooprofilattico Sperimentale delle Venezie), and D/bovine/France/5920/2014 (IDV) (from Dr. Mariette Ducatez, Université de Toulouse), all of which were propagated in MDCK cells.

Work using A/chicken/Scotland/054477/2021, an H5N1-2021 clade 2.3.4.4 HPAIV derived from a UK outbreak event and representative of the UK/European epizootic season in 2021, was carried out at SAPO containment level 4. The virus was propagated for two days in 9 to 10-day-old specified-pathogen-free embryonated eggs.

### Virus titration

For BSL-2 experiments, viral infectivity was quantified using plaque assay on MDCK cells. Virus samples were diluted in tissue culture medium to mitigate cytopathic effects associated with undiluted milk.

Plaques were visualised either by direct staining of the monolayer or, for IDV, by immunocytochemistry with a custom sheep polyclonal antibody against IDV NP (available from www.influenza.bio; third bleed, 1:500), followed by Alexa Fluor™ 568 donkey anti-sheep secondary antibody (Thermo, 1:1000) and a DAPI counterstain 1:500). Fluorescent signals were visualised using a Celigo imaging cytometer (Nexcelom).

For work at SAPO containment level 4, virus titration and assessment in milk was undertaken as described previously [8].

### Stability assay

Under BSL-2 conditions, virus stocks were diluted 1:10 (v/v) in test solutions. These were either buffered solutions (phosphate-buffered saline (PBS) or DMEM) or milk. Cow’s milk samples were either processed (homogenised and pasteurised milk, with whole milk (4% w/v fat), purchased from commercial suppliers in the UK, where no cases of bovine IAV had been confirmed at the time of study) or raw (obtained directly from cows in a herd managed by the University of Edinburgh, and used without prior processing). Pasteurised sheep’s milk was purchased from a commercial supplier in the UK. Milk was either used on the day of acquisition or kept refrigerated at 4⍰°C or frozen at −20⍰°C to prevent spoilage prior to experimentation. To test viral stability, diluted virus was aliquoted into (ThermoFisher), with sealed lids to prevent evaporation. These were placed in a sealed polystyrene box at room temperature or 4⍰°C and left for the indicated time.

For work at SAPO containment level 4, virus titration and assessment in milk was undertaken as described previously [8].

## Data Availability

The data needed to reproduce the findings and figures reported are available at the Open Science Framework (https://osf.io/gwezf/).

## Author Contributions

J.S. conceptualisation, methodology, investigation, data curation, writing – review and editing; C.J.W. conceptualisation, methodology, investigation, writing – review and editing; J.Y. conceptualisation, methodology, investigation, writing – review and editing; J.Z. conceptualisation, methodology, investigation, data curation, writing - original draft, writing – review and editing, visualization; S.J.C. methodology, investigation; J.C. investigation; K.D. investigation; N.McG. investigation; M.Q. conceptualisation, investigation; S.M.R investigation; N.P. investigation; W.S.T. investigation; S.K.W. investigation; A.C.B. writing—review and editing, supervision, funding acquisition; I.B. writing—review and editing, supervision, funding acquisition; P.D. conceptualisation, writing—review and editing, supervision, funding acquisition; M.I. writing—review and editing, supervision, funding acquisition; J.J. writing—review and editing, supervision, funding acquisition; T.P.P. resources, writing—review and editing, supervision, funding acquisition; E.H. conceptualisation, writing—original draft, writing—review and editing, supervision, funding acquisition.

## Acknowledgements

We thank Professor Alastair Macrae (Royal (Dick) School of Veterinary Science) for providing raw cows’ milk, Dr M Khalid Zakaria (MRC-University of Glasgow Centre for Virus Research) for assistance with preparing virus stocks, and the staff of the MRC Protein Phosphorylation and Ubiquitylation Unit, University of Dundee for their assistance in antibody generation. We gratefully acknowledge all data contributors, i.e., the authors and their originating laboratories responsible for obtaining the specimens, and their submitting laboratories for generating the genetic sequence and metadata and sharing via the GISAID Initiative, for the genomic data on which this research is based. All submitters of the data may be contacted directly via the GISAID website (https://www.gisaid.org).

## Supplementary Figures

**Fig S1.**
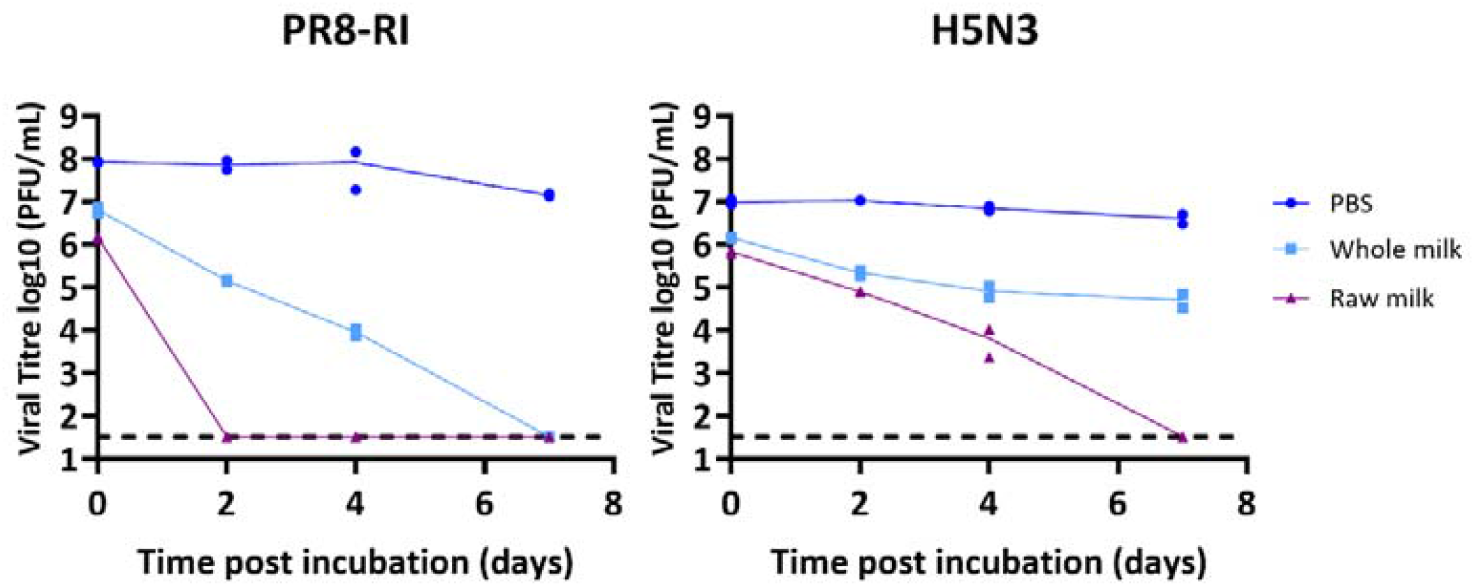
PR8 and H5N3 in raw milk and whole milk. Viruses were mixed with pasteurised whole milk or unpasteurised milk (raw milk), or PBS and either incubated at room temperature (∼20°C) or chilled (4°C) for the indicated times. Infectivity was measured by plaque assay. Data points show three independent repeats (each measured in duplicate), with lines connecting the mean values. Limit of detection (LoD) = 33 PFU/mL.

